# RCT-Twin-GAN Generates Digital Twins of Randomized Control Trials Adapted to Real-world Patients to Enhance their Inference and Application

**DOI:** 10.1101/2023.12.06.23299464

**Authors:** Phyllis M. Thangaraj, Sumukh Vasisht Shankar, Evangelos K. Oikonomou, Rohan Khera

**Author notes:** Both authors contributed equally (Phyllis M. Thangaraj, Sumukh Vasisht Shankar).

## Abstract

**Background:** Randomized clinical trials (RCTs) are designed to produce evidence in selected populations. Assessing their effects in the real-world is essential to change medical practice, however, key populations are historically underrepresented in the RCTs. We define an approach to simulate RCT-based effects in real-world settings using RCT digital twins reflecting the covariate patterns in an electronic health record (EHR).

**Methods:** We developed a Generative Adversarial Network (GAN) model, `RCT-Twin-GAN`, which generates a digital twin of an RCT (RCT-Twin) conditioned on covariate distributions from an EHR cohort. We improved upon a traditional tabular conditional GAN, `CTGAN`, with a loss function adapted for data distributions and by conditioning on multiple discrete and continuous covariates simultaneously. We assessed the similarity between a Heart Failure with preserved Ejection Fraction (HFpEF) RCT (TOPCAT), a Yale HFpEF EHR cohort, and RCT-Twin. We also evaluated cardiovascular event-free survival stratified by Spironolactone (treatment) use.

**Results:** By applying `RCT-Twin-GAN` to 3445 TOPCAT participants and conditioning on 3445 Yale EHR HFpEF patients, we generated RCT-Twin datasets between 1141-3445 patients in size, depending on covariate conditioning and model parameters. RCT-Twin randomly allocated spironolactone (S)/placebo (P) arms like an RCT, was similar to RCT by a multi-dimensional distance metric, and balanced covariates (median absolute standardized mean difference (MASMD) 0.017, IQR 0.0034-0.030). The 5 EHR-conditioned covariates in RCT-Twin were closer to the EHR compared with the RCT (MASMD 0.008 vs 0.63, IQR 0.005-0.018 vs 0.59-1.11). RCT-Twin reproduced the overall effect size seen in TOPCAT (5-year cardiovascular composite outcome odds ratio (95% confidence interval) of 0.89 (0.75-1.06) in RCT vs 0.85 (0.69-1.04) in RCT-Twin).

**Conclusions:** `RCT-Twin-GAN` simulates RCT-derived effects in real-world patients by translating these effects to the covariate distributions of EHR patients. This key methodological advance may enable the direct translation of RCT-derived effects into real-world patient populations and may enable causal inference in real-world settings.

## 1. Introduction

Randomized control trials (RCTs) are the gold standard for identifying practice-changing management of disease, but their generalizability to real-world patients remains a challenge. Underrepresentation of key patient demographic and comorbid groups contributes to this gap, resulting in difficulty applying treatment effect results from RCTs to underrepresented patient populations (Patel et al., 2017; Lim et al., 2022). One of several large-scale examples, the Treatment of Preserved Cardiac Function Heart Failure with an Aldosterone Antagonist Trial (TOPCAT), demonstrated heterogeneous treatment effects across participants, questioning the extent to which the trial cohort is representative of the real-world (Pitt et al., 2014; Pfeffer et al., 2015; Cohen et al., 2020). Nevertheless, in addition to the randomization of tested interventions, RCTs enable a comprehensive phenotypic evaluation of the baseline composition of a population. An equivalent cohort more representative of real-world patients is needed to better inform treatment efficacy and identify patients who should be targeted for future RCTs. The electronic health record (EHR) is a promising research resource for extracting clinical and sociodemographic information about real-world patients but is limited by lack of use of novel treatments evaluated in RCTs, non-random missingness of data, ascertainment bias, and infrequent follow-up (Dhingra et al., 2023).

Generative Adversarial Networks (GANs) can amalgamate RCT and EHR cohort characteristics by building a synthetic cohort representative of their covariate distributions. Several advances have been made to GANs to address the challenges of EHR data (Ghosheh et al., 2022). We developed `RCT-Twin-GAN`, which builds a synthetic twin RCT dataset reflective of a real-world patient population by conditioning on covariates from an equivalent EHR population. We show more optimal alignment between the digital twin cohort and EHR cohort compared to other models, `CTGAN` and `EHR-M-GAN` (Xu et al., 2019; Li et al., 2023). We also show successful random allocation of treatment arms similar to RCT while also improving the representation of key patient populations by incorporating EHR patient covariate distributions. We expect true effect estimates and variance in a disease population to be reflective of all patients evaluated and/or treated for the disease. Our model contributes to the evidence for creating an RCT digital twin more reflective of real-world populations for enhancing inference for real-world patients.

## 2. Related work

Although there have been deep generative models emulating treatment effects in RCTs, they have not considered the under-enrollment of key patient populations such as women, minorities, the elderly, and patients with many comorbidities (Liu et al., 2021; Ge et al., 2020; Yoon et al., 2018). Other studies have tried to estimate treatment effects in EHR patient populations, which is plagued by confounding due to lack of randomization (Chu et al., 2020). (Averitt et al., 2020) developed the Counterfactual *χ*`-GAN`, which used Pearson divergence to balance covariates between two observational study populations. Conditional Tabular GAN (`CTGAN`) successfully built synthetic datasets of mixed-type tabular data (Xu et al., 2019). They developed advances such as mode-specific normalization to better model the distribution of continuous data, a conditional generator, and training-by-sampling to handle highly imbalanced covariates. We built upon `CTGAN` to create `RCT-Twin-GAN` that efficiently handles multiple mixed data types to generate a digital twin of an RCT cohort conditioned on EHR covariates. We compare the cohort similarity performance of our model with `CTGAN` and `EHR-M-GAN`, which pretrains using a dual-variational autoencoder (VAE) that captures mixed-type time-series EHR data (Xu et al., 2019; Li et al., 2023).

## 3. Methods

### 3.1. Study Cohorts and Covariates

We extracted patient-level information from RCT and EHR data sources. The first cohort, TOPCAT, was a multi-center international RCT that recruited 3,445 subjects with Heart Failure with Preserved Ejection Fraction (HFpEF) and studied the effect of spironolactone on the incidence of death from cardiovascular cause, myocardial infarction, stroke, aborted cardiac arrest, and hospitalization for decompensated heart failure. See Appendix A for further details. The second cohort included 10,467 patients admitted with heart failure in the Yale New Haven Health System (YNHHS), and imaging confirmed HFpEF. We extracted baseline demographics, conditions, procedures, vital signs, medications, laboratory values, and imaging from the TOPCAT participant dataset. We chose 65 variables, 39 with discrete and 24 with continuous data across these categories to map to the EHR. Pre-processing of data is described in Appendix (B). The study was reviewed by Yale IRB and deemed exempt as it uses retrospective data.

### 3.2. RCT-Twin-GAN Model

We modeled `RCT-Twin-GAN` after `CTGAN`, which uses a mode-specific normalization to overcome covariate non-Gaussian multimodal distribution and a conditional generator and training by sampling approach to handle imbalanced covariates. We train `RCT-Twin-GAN` on the 65 RCT covariates, treatment arm, and outcomes. `RCT-Twin-GAN` updates the cross-entropy loss function used in `CTGAN` with Mean Absolute Error (MAE) to enable computationally efficient generation of EHR-conditioned RCT with more continuous variables.

As in `CTGAN`, `RCT-Twin-GAN` used a rectified linear unit (ReLU) activation function and Gumbel Soft-max in the generator and a leaky ReLU function and dropout in the discriminator. We trained with an ADAM Optimizer and a learning rate of 10^*−*4^. Other modifications included training for 500 epochs and a batch size of 128. For conditional sampling, we used a batch size of 32 with 500 maximum tries per batch to minimize the number of rows that could not be produced. During the runtime analysis, we reduced the batch size to 1 with 200 maximum tries to maximize speed.

We incorporated the Python library from Synthetic Data Vault (SDV) to condition on multiple known columns and sample the remaining required columns using Conditional Parameter Aggregation (CPA) (Patki et al., 2016). We also make use of conditioning on both continuous and discrete covariate distributions from SDV. This is a modification from `CTGAN`, which used a reject sampling approach to condition on one column at a time and only on discrete data. The generative model for tabular data in this method consists of distributions, which describe a value in a column using a Gaussian or uniform distribution, and covariances, which describe the correlation between values in the same row using the multivariant Gaussian Copula (Patki et al., 2016). This model converts all column distributions to standard normal before finding the covariances. When *m* columns of a dataset of *n* columns are passed as conditional columns to the generator, each row of the conditional columns is considered as a vector - thus having a vector size of *m*. For each such vector, using the covariances and CPA, new *n − m* columns for the vector are sampled. Once such a row is created, it is appended to the sampled data frame to the corresponding conditional vector. `RCT-Twin-GAN` completes this sampling process by using covariate distributions found in the EHR population. Figure 1 outlines the cohort generation process and training and conditioning of `RCT-Twin-GAN`.

**Figure 1:**
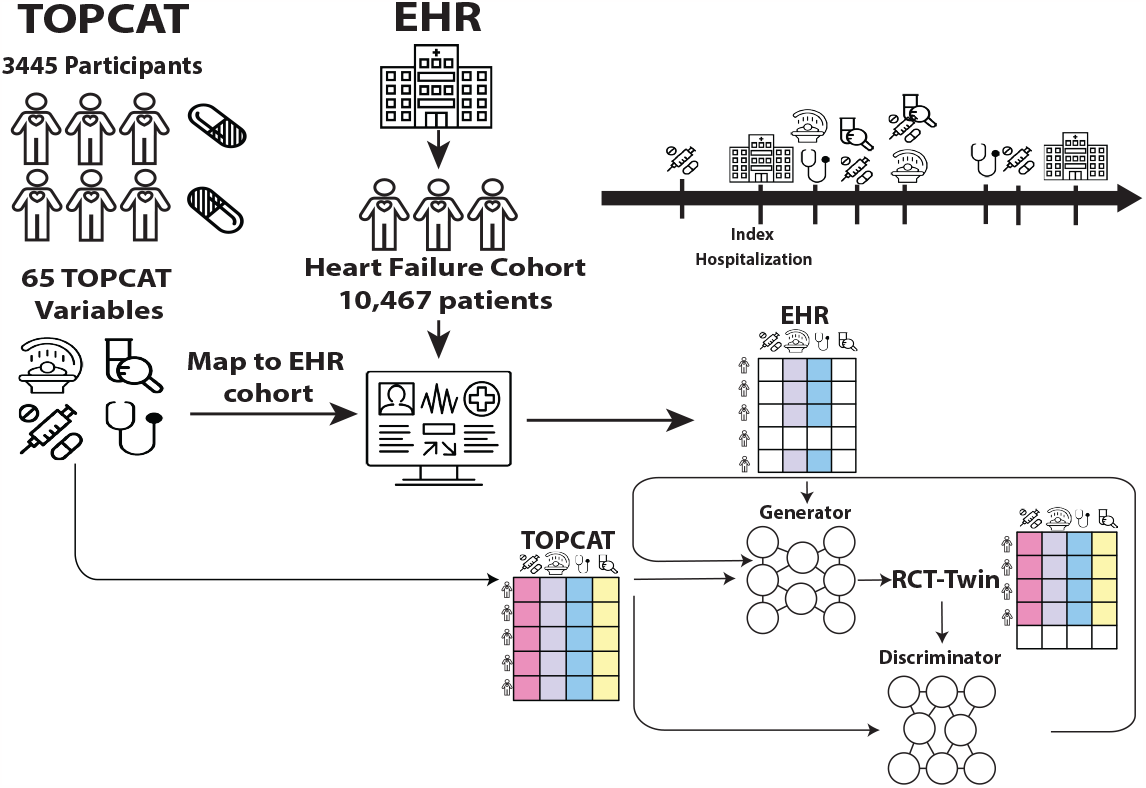
Graphical depiction of RCT and EHR cohort generation and development of `RCT-Twin-GAN`. The top depicts the curation of the RCT participants from each treatment arm and their covariates, the curation of the EHR heart failure cohort from the index hospitalization for heart failure, and the mapping of the RCT covariates to EHR variables. The bottom shows the training of the GAN generator with the RCT cohort and conditioning with select equivalent covariates from the EHR cohort, creating the RCT-Twin dataset. The discriminator then evaluates the similarity of RCT-Twin with the original RCT data and feeds its evaluation back to the generator.

We analyzed the balance, similarity, and cardiovascular outcomes of three different synthetic datasets. First, the RCT twin, in which `RCT-Twin-GAN` is trained on the RCT cohort without any conditioning. The next synthetic dataset was trained on the RCT cohort but conditioned on 5 covariates from the RCT cohort. The final synthetic dataset was also trained on the RCT cohort but conditioned on the same 5 covariates from the EHR cohort. We conditioned with the top 5 covariates correlated with the composite outcome in the RCT cohort placebo treatment arm. These were age, left ventricular ejection fraction (EF), heart rate, the laboratory value creatinine, and whether the patient used the medication class ACE Inhibitors. We chose these covariates as a representative example. Accordingly, any continuous and discrete covariates can be used by the model.

We also completed a runtime and synthetic cohort generation analysis comparing 10 different conditioning parameters. We also changed the synthetic data generation model to create one row at a time rather than a batch at a time to improve speed. First, the RCT twin, in which `RCT-Twin-GAN` is trained on the RCT cohort without any conditioning. The next synthetic dataset was trained on the RCT cohort but conditioned on 1, 3, 5, and 7 continuous covariates from the RCT cohort. The final synthetic dataset was also trained on 3 discrete covariate distributions from the RCT cohort. Similarly, synthetic datasets were trained on the RCT cohort and conditioned on the same 1,3,5, and 7 continuous and three discrete covariates, but this time the distributions were from the EHR cohort. We conditioned with the top covariates correlated with the composite outcome in the RCT cohort placebo treatment arm. The continuous covariates started with just creatinine, and then ejection fraction, heart rate, age, systolic blood pressure, potassium, BMI, gender, whether the patient was on an ACE inhibitor or ARB, and whether the patient had dyslipidemia were added sequentially. The final synthetic dataset was trained on the RCT cohort and conditioned on 10 random covariate distributions from the EHR cohort.

### 3.3. Statistical Analysis

We quantified the difference between two cohorts by calculating Gower’s dissimilarity distance between individuals within each cohort. We chose the Gower distance because of its ability to handle multi-dimensional and mixed-type data, specifically categorical and continuous. Rather than compare single covariates at a time, this distance is a complex representation of all covariates. The Gower’s distance was calculated as described in (Gower, 1971). Briefly, continuous variable distances were calculated as the absolute value difference divided by range, and binary variable distance was assigned 1 if identical and 0 otherwise. The distance per patient was then calculated as the average across all variables. We calculated the mean of median Gower’s distances between each cohort participant. We also visualized the distances between cohort participants with a dimensionality reduction method called uniform manifold approximation and projection (UMAP) (McInnes et al., 2018).

We evaluated the similarity of covariate distribution of RCT-Twin to the RCT cohort and EHR cohort by calculating the median absolute standardized mean difference of covariates between the different cohorts/datasets.

#### 3.3.1. Correlation between Covariates

We assessed the similarity of covariate correlation between the real and synthetic data sets by measuring the mean absolute difference (MAD) of Spearman correlation coefficients (SCCs) between five different cohorts: (1) the RCT cohort, (2) the RCT Twin, (3) the RCT Twin conditioned on RCT cohort, (4) the RCT Twin conditioned on EHR cohort, and (5) the EHR cohort, based on prior work (Li et al., 2023). Spearman correlation rather than Pearson correlation permitted use in mixed-type datasets without presumed normality of covariate distribution. We also measured CorAcc, in which we discretized the SCCs according to the bins described in (Li et al., 2023) and calculated the percentage of discretized SCCs that are equal between the two cohorts.

We also calculated the 5-year odds ratio of composite cardiovascular death, cardiac arrest, and hospitalization for heart failure to evaluate RCT-Twin-GAN’s ability to emulate RCT treatment effects.

## 4. Results

### 4.1. Gower’s Dissimilarity and Spearman’s Correlation between cohorts

All real and synthetic datasets were separated by treatment arm (spironolactone (S) or placebo (P)). The median (sem) of the median Gower’s distance between the RCT S. Arm cohort and the other cohorts ranged from 0.189 (0.000686) in the RCT P. Arm to 0.213 (0.000760) in the EHR. (Figure 2(*b*)). The EHR was furthest from the RCT S. Arm, and out of the synthetic datasets, the RCT-Twin dataset conditioned on EHR data was furthest (0.198 (.000645)). A UMAP of the Gower’s distance between individuals in each cohort showed separation between the original EHR and all other cohorts(Figure 2).

**Figure 2:**
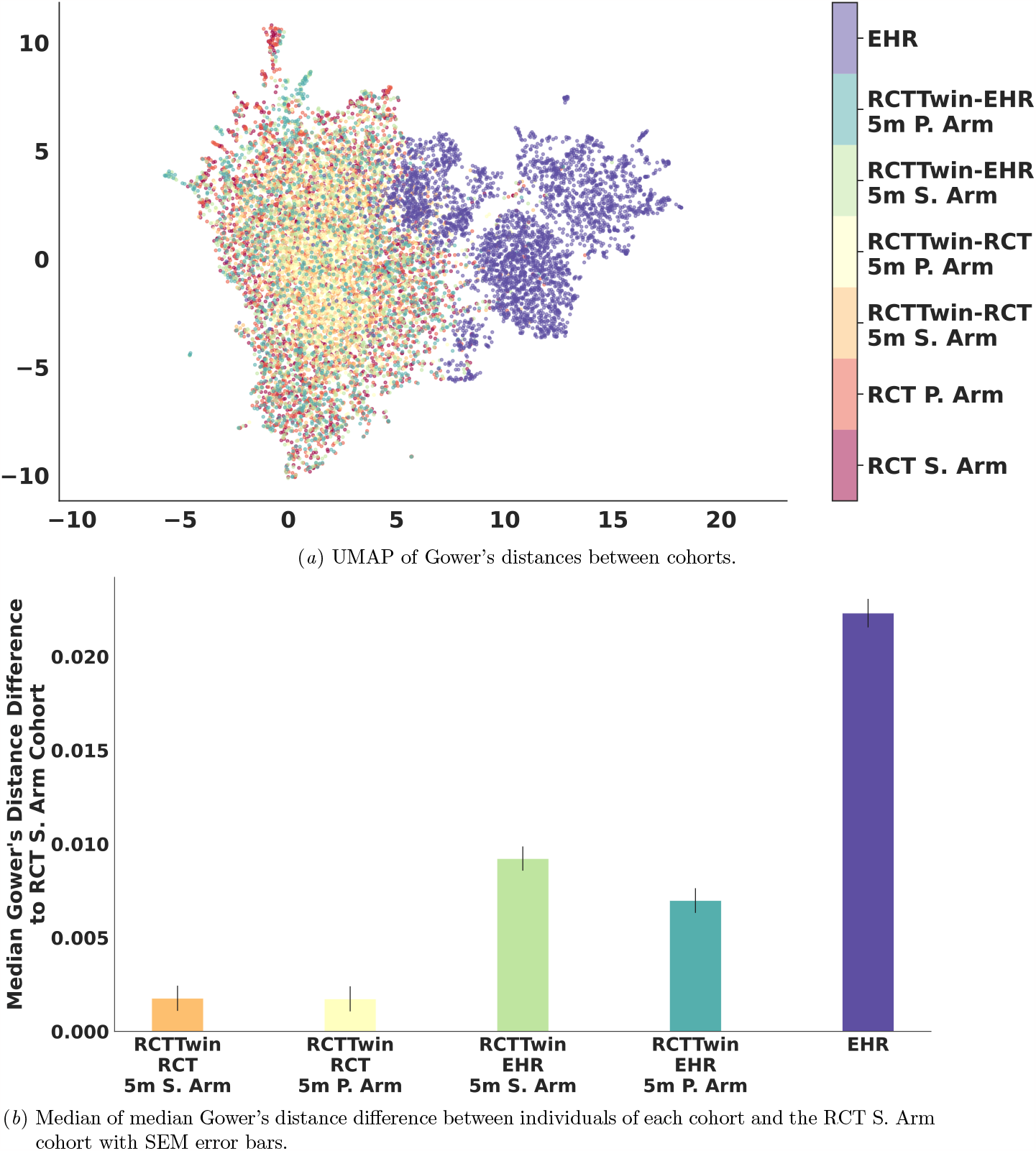
The X and Y axes in (a) are UMAP coordinates. Each color represents a cohort. Orange is the RCT Placebo (P.) arm, and Red is the RCT Spironolactone (S.) arm. Yellow is the RCT-Twin placebo cohort conditioned on 5 covariates using the RCT data itself (placebo label generated by the model) (RCT-Twin-RCT 5m P. Arm), Light Orange is the RCT-Twin spironolactone Arm conditioned on the same 5 RCT covariates (spironolactone label generated by the model) (RCT-Twin-RCT 5m S. Arm), B6lue is the RCT-Twin placebo arm conditioned on the same 5 covariates drawn from EHR cohorts (placebo label generated by the model) (RCT-Twin-EHR 5m P. Arm), Green is the RCT-Twin spironolactone arm conditioned on the same EHR data (spironolactone label generated by the model) (RCT-Twin-EHR 5m S. Arm), and Purple is the EHR cohort. (B) The Y axis is the median of median pairwise Gower’s distance between the labeled cohort and RCT spironolactone arm, referenced to the distance between the spironolactone and placebo arms of the RCT.

The lowest mean absolute difference (MAD) of Spearman’s Correlation coefficients (SCC), or highest correlation, between covariates of two cohorts resulted from the `RCT-Twin-GAN` model compared to `CTGAN` and `EHR-M-GAN`. Within the cohorts, the synthetic data sets were most correlated to each other (MAD 0.0184,0.0274, and 0.0290). The lowest MAD of SCC with a real dataset included the EHR cohort and RCT Twin conditioned on EHR data, 0.0592. Correspondingly, the synthetic data sets had the highest CorrAcc (98.3%-98.9%) between them, and the RCT Twin conditioned on RCT or EHR data both had the next highest CorrACC with the EHR cohort (82.9%). `CTGAN` had less similarity between the RCT Twin conditioned on EHR data and EHR (MAD 0.0621, CorACC 82.4%). The `EHR-M-GAN` model had high MAD of SCCs (median 0.165, IQR 0.0789-0.190) and low CorrAcc (median 43.0%, IQR 32.0%-63.0%) (Table 1).

**Table 1:**
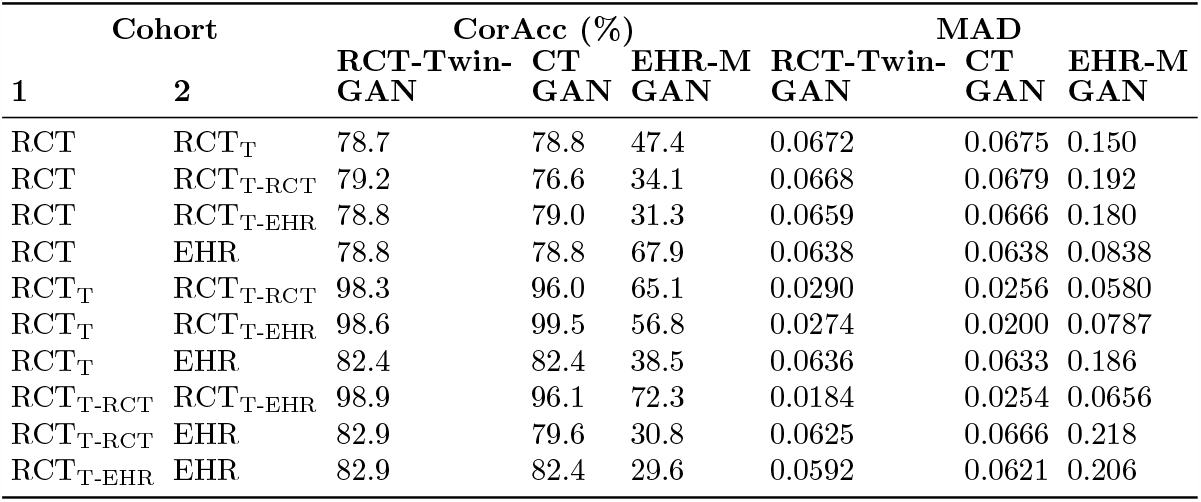
Mean absolute difference (MAD) and CorAcc of Spearman Correlation Coefficients between cohorts. RCT_T_ refers to RCT Twin cohort, RCT_T-RCT_ refers to the RCT Twin conditioned on RCT data cohort, and RCT_T-EHR_ refers to the RCT Twin conditioned on EHR data cohort.

| Cohort |  | CorAcc (%) |  |  | MAD |  |  |
| --- | --- | --- | --- | --- | --- | --- | --- |
| 1 | 2 | RCT-Twin-GAN | CT GAN | EHR-M GAN | RCT-Twin-GAN | CT GAN | EHR-M GAN |
| RCT | $RCT_T$ | 78.7 | 78.8 | 47.4 | 0.0672 | 0.0675 | 0.150 |
| RCT | $RCT_{T-RCT}$ | 79.2 | 76.6 | 34.1 | 0.0668 | 0.0679 | 0.192 |
| RCT | $RCT_{T-EHR}$ | 78.8 | 79.0 | 31.3 | 0.0659 | 0.0666 | 0.180 |
| RCT | EHR | 78.8 | 78.8 | 67.9 | 0.0638 | 0.0638 | 0.0838 |
| $RCT_T$ | $RCT_{T-RCT}$ | 98.3 | 96.0 | 65.1 | 0.0290 | 0.0256 | 0.0580 |
| $RCT_T$ | $RCT_{T-EHR}$ | 98.6 | 99.5 | 56.8 | 0.0274 | 0.0200 | 0.0787 |
| $RCT_T$ | EHR | 82.4 | 82.4 | 38.5 | 0.0636 | 0.0633 | 0.186 |
| $RCT_{T-RCT}$ | $RCT_{T-EHR}$ | 98.9 | 96.1 | 72.3 | 0.0184 | 0.0254 | 0.0656 |
| $RCT_{T-RCT}$ | EHR | 82.9 | 79.6 | 30.8 | 0.0625 | 0.0666 | 0.218 |
| $RCT_{T-EHR}$ | EHR | 82.9 | 82.4 | 29.6 | 0.0592 | 0.0621 | 0.206 |

### 4.2. Mean Standardized Difference between cohorts and digital twin

By applying RCT-Twin-GAN to 3445 TOPCAT participants and conditioning on 3445 Yale EHR HFpEF patients, we generated a 2173-patient RCT-Twin. RCT-Twin randomly allocated spironolactone (S)/Placebo (P) arms like RCT, were similar to RCT by a multi-dimensional distance metric and balanced covariates (median absolute standardized mean difference (MASMD) 0.017, IQR 0.0034-0.030) (Figure 3). The 5 EHR-conditioned covariates in RCT-Twin were closer to the EHR compared to RCT (MASMD 0.008 vs 0.63, IQR 0.005-0.018 vs 0.59-1.11) (Figure 3(*b*)).

**Figure 3:**
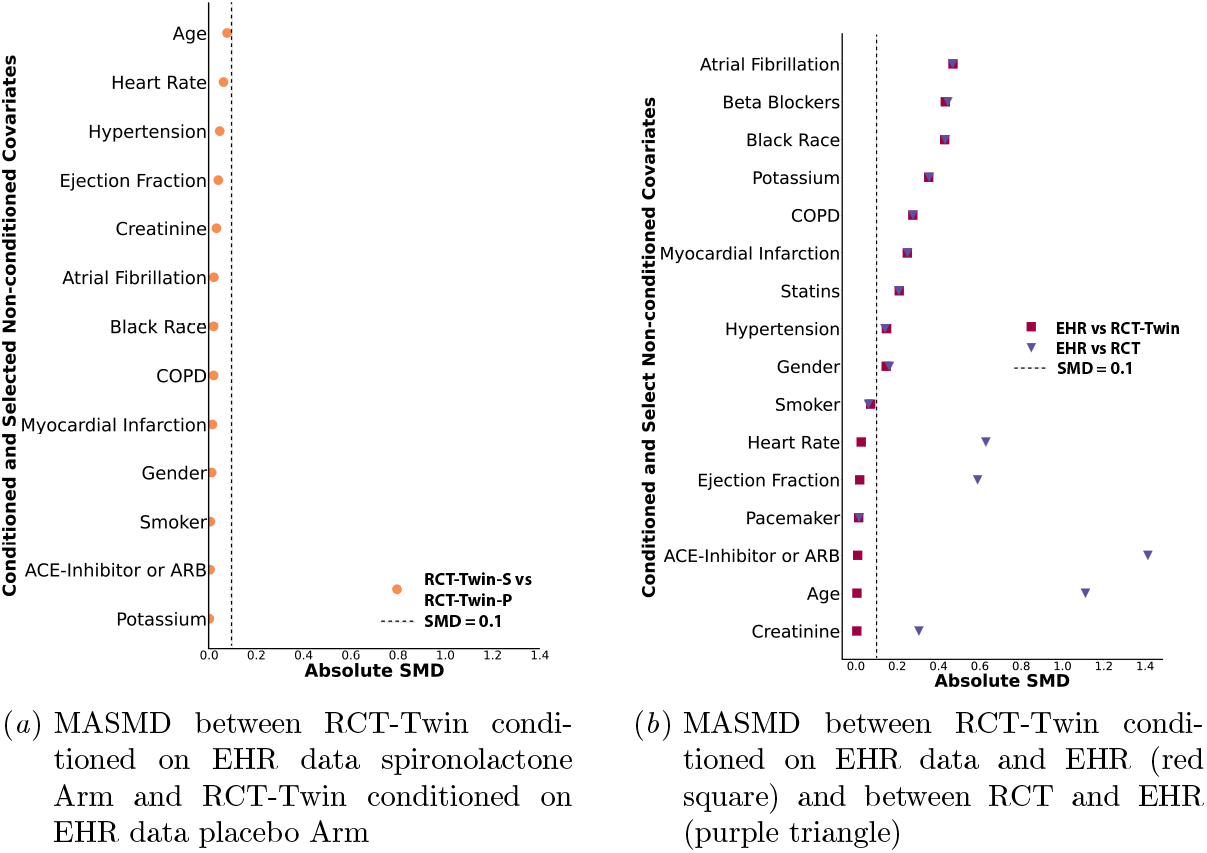
Median absolute standardized mean difference (MASMD) between (Left) RCT-Twin conditioned on EHR data Spironolactone Arm (RCT-Twin-S) and RCT-Twin conditioned on EHR data Placebo Arm (RCT-Twin-P) and (Right) EHR cohort and RCT-Twin conditioned on EHR data (red square) and EHR and RCT (purple triangle). Conditioning columns were age, EF, heart rate, creatinine, and an ACE-inhibitor. The dotted line represents an MASMD of 0.1.

### 4.3. Survival Analysis Comparison between RCT-Twin and original RCT

The RCT-Twin dataset had similar outcomes to the RCT cohort in that there was no difference in survival across treatment arms. RCT 5-year odds ratio of cardiovascular death, cardiac arrest, or hospitalization for heart failure was 0.89, 95% confidence interval (CI) 0.75-1.06 while RCT-Twin 5-year odds ratio of the same outcome was 0.85, 95% CI 0.69-1.04.

### 4.4. Run time of models

We conditioned `RCT-Twin-GAN` on 5 discrete columns due to increased run time and memory usage with continuous columns. Generating 500 rows of synthetic data took 947 seconds when conditioning with 5 discrete columns, 8,023 seconds when conditioning with 10 discrete columns, 10,730 seconds when conditioning with 5 continuous columns, and 25,610 seconds when conditioning with 10 continuous columns. Table 2 shows our runtime analysis across 10 different combinations of covariate distribution and data conditioning. The more covariates conditioned on, the longer the run time and fewer rows generated. Conditioning on EHR data took a longer time than RCT data.

**Table 2:** Comparison of Run times and number of synthetic rows generated. The first column describes the conditioning data, the second column describes the number (No.) of continuous (cont.) columns (col) on which RCT-Twin-GAN was conditioned, the third column describes the number of discrete (disc.) columns on which RCT-Twin-GAN was conditioned, the fourth column is the run time in seconds, and the fifth column is the number of synthetic rows generated by RCT-Twin-GAN.

| Conditioning Data | No. Cont. Col. | No. Disc. Col. | Run Time (s) | No. Rows Generated |
| --- | --- | --- | --- | --- |
| RCT | 1 | 0 | 324.6 | 3445 |
| EHR | 1 | 0 | 726.4 | 3445 |
| RCT | 3 | 0 | 575.2 | 3444 |
| EHR | 3 | 0 | 2061 | 3339 |
| RCT | 5 | 0 | 957.3 | 3433 |
| EHR | 5 | 0 | 4680 | 3008 |
| RCT | 7 | 3 | 5584 | 3078 |
| EHR | 7 | 3 | 17660 | 2279 |
| EHR | 0 | 10 | 3256 | 2960 |

**Table 3:** Abbreviations within the Paper.

| Variable | Abbreviation |
| --- | --- |
| EHR | Electronic Health Record |
| RCT | Randomized Clinical Trial |
| No. | Number |
| Cont. | Continuous |
| Col. | Column |
| Disc. | Discrete |
| S. Arm | Spironolactone Arm |
| P. Arm | Placebo Arm |
| RCT-RCT-Twin 1c | RCT-Twin dataset, GAN conditioned on one continuous covariate of RCT |
| RCT-RCT-Twin 3c | RCT-Twin dataset, GAN conditioned on three continuous covariates of RCT |
| RCT-RCT-Twin 5c | RCT-Twin dataset, GAN conditioned on five continuous covariates of RCT |
| RCT-RCT-Twin 7c3d | RCT-Twin dataset, GAN conditioned on seven continuous and three discrete covariates of RCT |
| RCT-Twin-EHR 1c | RCT-Twin dataset, GAN conditioned on one continuous covariate of EHR |
| RCT-Twin-EHR 3c | RCT-Twin dataset, GAN conditioned on three continuous covariates of EHR |
| RCT-Twin-EHR 5c | RCT-Twin dataset, GAN conditioned on five continuous covariates of EHR |
| RCT-Twin-EHR 7c3d | RCT-Twin dataset, GAN conditioned on seven continuous and three discrete covariates of EHR |
| RCT-RCT-Twin 5m | RCT-Twin dataset, GAN conditioned on 5 mixed (4 cont., 1 disc.) covariates of RCT |
| RCT-Twin-EHR 5m | RCT-Twin dataset, GAN conditioned on 5 mixed (4 cont., 1 disc.) covariates of EHR |
| HFpEF | Heart Failure with Preserved Ejection Fraction |

## 5. Discussion

Participants of RCTs do not always represent real-world patients, making the generalizability of gold standard evidence a challenge. In this study, we developed `RCT-Twin-GAN`, a conditional generative adversarial model that creates a synthetic RCT cohort reflective of a real-world patient population. Using the heart failure RCT TOPCAT as an example, we create a synthetic cohort more similar to an EHR cohort than TOPCAT when conditioning with EHR data. We showed this by demonstrating a similar distribution of RCT-Twin to the EHR in the EHR-conditioned covariates. We also show that RCT-Twin randomly allocates treatment arms as seen in an RCT cohort and similarly showed no survival benefit in the spironolactone treatment arm as seen in the original TOPCAT trial. We also show a higher covariate correlation between the synthetic cohort and EHR cohort compared to two other models. We have introduced a new application of GANs to build synthetic cohorts by creating an RCT digital twin reflective of real-world patients found in the EHR. Our study demonstrates a way to evaluate the generalizability of an RCT to the general population by embedding covariate distributions that are more representative of real-world populations. This amplifies the effects for those who are more frequently seen in clinical practice.

There are limitations to consider. First, there is no perfect representation of real-world patients. The EHR patients seeking hospital care likely represent a sicker subset of the HFpEF population compared to the TOPCAT cohort, highlighting an important cross-section of eligible patients. Second, we only conditioned on five out of a possible 65 variables due to run time length. Conditioning on a higher number of continuous variables could improve model performance, but run-time efficiency will need to be addressed. In addition, the Gower distances of the RCT-Twin dataset from the EHR and RCT cohorts were higher than those datasets conditioned on covariates, which we attribute to a sampling variation. RCTs enroll from the disease population; therefore, they represent samples from the overall population.

We introduce `RCT-Twin-GAN` as a key methodological advance to enable the direct translation of RCT-derived effects into real-world patient populations.

## 6. Acknowledgements

This study will be presented at ML4H 2023.

## Funding

The study is supported by grants from the National Heart, Lung, and Blood Institute (NHLBI) to RK (K23HL153775), PMT (5T32HL155000-03), and EKO (1F32HL170592-01).

## Disclosures

The authors are coinventors of a provisional patent related to the current work (63/606,203). EKO is a co-inventor of the U.S. Patent Applications 63/508,315 63/177,117, a cofounder of Evidence2Health (with RK), and has previously served as a consultant to Caristo Diagnostics Ltd (outside the present work). RK is an Associate Editor of JAMA. He receives support from the Doris Duke Charitable Foundation (under award, 2022060). He also receives research support, through Yale, from Bristol-Myers Squibb, Novo Nordisk, and BridgeBio. He is a coinventor of U.S. Provisional Patent Applications 63/177,117, 63/428,569, 63/346,610, 63/484,426, 63/508,315, and 63/606,203 and is a co-founder of Ensight-AI and Evidence2Health, health platforms to improve cardiovascular diagnosis and evidence-based cardiovascular care.

## 7. Data Availability

The TOPCAT data are available online (https://biolincc.nhlbi.nih.gov/studies/topcat/), and the EHR data is not available to maintain patient privacy.

## Appendix A. Details of Study Cohorts

Details of TOPCAT (ClinicalTrials.gov identifier: NCT00094302) have been previously published (Pitt et al., 2014). The study enrolled 3445 individuals 50 years globally with left ventricular ejection fraction (LVEF) 45%, one sign and one symptom of heart failure in the last 12 months, and at least one hospitalization for heart failure in the last 12 months. Participants were randomized to receive either spironolactone or placebo.

## Appendix B. Imputation of missing variables

We imputed missing values by running the MissForest package from missingpy in Python, which chains random forests to predict imputed values that minimize the root mean square between complete and imputed data. This algorithm is the most effective imputation algorithm in mixed-type categorical and continuous data and high-dimensional data(Stekhoven and Bühlmann, 2012; noa).

## Appendix C. Abbreviations

## Notes

### Funding Statement

This study was funded by:
K23HL153775
5T32HL155000-03
1F32HL170592-01

### Author Declarations

IRB of Yale University waived ethical approval and granted a waiver of consent for this work since this was a retrospective study with minimal risks to subjects.

### Summary of Updates

We have revised the name of the GAN to RCT-Twin-GAN.

## Citations and Bibliography References

MissForest. https://pypi.org/project/MissForest/. Accessed: 2023-4-2.

Amelia J Averitt, Natnicha Vanitchanant, Rajesh Ranganath, and Adler J Perotte. The counterfactual χ-GAN: Finding comparable cohorts in observational health data. J. Biomed. Inform., 109: 103515, September 2020.

Jiebin Chu, Wei Dong, Jinliang Wang, Kunlun He, and Zhengxing Huang. Treatment effect prediction with adversarial deep learning using electronic health records. BMC Med. Inform. Decis. Mak., 20 (Suppl 4):139, December 2020.

Jordana B Cohen, Sarah J Schrauben, Lei Zhao, Michael D Basso, Mary Ellen Cvijic, Zhuyin Li, Melissa Yarde, Zhaoqing Wang, Priyanka T Bhattacharya, Diana A Chirinos, Stuart Prenner, Payman Zamani, Dietmar A Seiffert, Bruce D Car, David A Gordon, Kenneth Margulies, Thomas Cappola, and Julio A Chirinos. Clinical phenogroups in heart failure with preserved ejection fraction: Detailed phenotypes, prognosis, and response to spironolactone. JACC. Heart failure, 8 (3):172–184, March 2020.

Lovedeep Singh Dhingra, Miles Shen, Anjali Mangla, and Rohan Khera. Cardiovascular care innovation through Data-Driven discoveries in the electronic health record. Am. J. Cardiol., 203:136–148, July 2023.

Qiyang Ge, Xuelin Huang, Shenying Fang, Shicheng Guo, Yuanyuan Liu, Wei Lin, and Momiao Xiong. Conditional generative adversarial networks for individualized treatment effect estimation and treatment selection. Front. Genet., 11:585804, December 2020.

Ghadeer Ghosheh, Jin Li, and Tingting Zhu. A review of generative adversarial networks for electronic health records: applications, evaluation measures and data sources. March 2022.

J C Gower. A general coefficient of similarity and some of its properties. Biometrics, 27(4):857–871, 1971.

Jin Li, Benjamin J Cairns, Jingsong Li, and Tingting Zhu. Generating synthetic mixed-type longitudinal electronic health records for artificial intelligent applications. NPJ Digit Med, 6(1):98, May 2023.

Yvonne Mei Fong Lim, Megan Molnar, Ilonca Vaartjes, Gianluigi Savarese, Marinus J C Eijkemans, Alicia Uijl, Eleni Vradi, Kiliana Suzart-Woischnik, Jasper J Brugts, Hans-Peter Brunner-La Rocca, Vanessa Blanc-Guillemaud, Fabrice Couvelard, Claire Baudier, Tomasz Dyszynski, Sandra Waechter, Lars H Lund, Arno W Hoes, Benoit Tyl, Folkert W Asselbergs, Christoph Gerlinger, Diederick E Grobbee, Maureen Cronin, and Stefan Koudstaal. Generalizability of randomized controlled trials in heart failure with reduced ejection fraction. Eur Heart J Qual Care Clin Outcomes, 8(7):761–769, October 2022.

Ruishan Liu, Shemra Rizzo, Samuel Whipple, Navdeep Pal, Arturo Lopez Pineda, Michael Lu, Brandon Arnieri, Ying Lu, William Capra, Ryan Copping, and James Zou. Evaluating eligibility criteria of oncology trials using real-world data and AI. Nature, 592(7855):629–633, April 2021.

Leland McInnes, John Healy, and James Melville. UMAP: Uniform manifold approximation and projection for dimension reduction. February 2018.

Hitesh C Patel, Carl Hayward, Jason N Dungu, Sofia Papadopoulou, Abdel Saidmeerasah, Robin Ray, Carlo Di Mario, Nesan Shanmugam, Martin R Cowie, and Lisa J Anderson. Assessing the eligibility criteria in phase III randomized controlled trials of drug therapy in heart failure with preserved ejection fraction: The critical Play-Off between a “pure” patient phenotype and the generalizability of trial findings. J. Card. Fail., 23(7):517–524, July 2017.

Neha Patki, Roy Wedge, and Kalyan Veeramachaneni. The synthetic data vault. In 2016 IEEE International Conference on Data Science and Advanced Analytics (DSAA), pages 399–410, October 2016.

Marc A Pfeffer, Brian Claggett, Susan F Assmann, Robin Boineau, Inder S Anand, Nadine Clausell, Akshay S Desai, Rafael Diaz, Jerome L Fleg, Ivan Gordeev, John F Heitner, Eldrin F Lewis, Eileen O’Meara, Jean-Lucien Rouleau, Jeffrey L Probstfield, Tamaz Shaburishvili, Sanjiv J Shah, Scott D Solomon, Nancy K Sweitzer, Sonja M McKinlay, and Bertram Pitt. Regional variation in patients and outcomes in the treatment of preserved cardiac function heart failure with an aldosterone antagonist (TOPCAT) trial. Circulation, 131(1):34–42, January 2015.

Bertram Pitt, Marc A Pfeffer, Susan F Assmann, Robin Boineau, Inder S Anand, Brian Claggett, Nadine Clausell, Akshay S Desai, Rafael Diaz, Jerome L Fleg, Ivan Gordeev, Brian Harty, John F Heitner, Christopher T Kenwood, Eldrin F Lewis, Eileen O’Meara, Jeffrey L Probstfield, Tamaz Shaburishvili, Sanjiv J Shah, Scott D Solomon, Nancy K Sweitzer, Song Yang, and Sonja M McKinlay. Spironolactone for heart failure with preserved ejection fraction. N. Engl. J. Med., 370 (15):1383–1392, April 2014.

Daniel J Stekhoven and Peter Bühlmann. MissForest–non-parametric missing value imputation for mixed-type data. Bioinformatics, 28 (1):112–118, January 2012.

Lei Xu, Maria Skoularidou, Alfredo Cuesta-Infante, and Kalyan Veeramachaneni. Modeling tabular data using conditional GAN. June 2019.

Jinsung Yoon, James Jordon, and Mihaela Van Der Schaar. Ganite: Estimation of individualized treatment effects using generative adversarial nets. https://openreview.net/pdf?id=ByKWUeWA-, 2018. Accessed: 2023-11-9.

